# Derivation and validation of indices incorporating vasopressor dose and blood pressure values over time

**DOI:** 10.1101/2024.09.21.24313905

**Authors:** Alain Gervais, François Lamontagne, Jean-Baptiste Michaud, Neill KJ Adhikari, Jean-Michel Pagé, Marie-Hélène Masse, Michael O Harhay, Michaël Chassé, Félix Lamontagne, Katia Laforge, Alexandra Fortin, Marc-André Leclair, Simon Lévesque, Marie-Pier Domingue, Neda Momenzadeh, Martin Vallières, Ruxandra Pinto, Maxime Morin-Lavoie, Francis Carter, Félix Camirand Lemyre

**Affiliations:** Soins intensifs de l’hôpital Santa-Cabrini Ospedale, Montréal, Québec, Canada; Université de Montréal, Montréal, Québec, Canada; Faculté de médecine et des sciences de la santé, Université de Sherbrooke, Sherbrooke, Québec, Canada; Centre de Recherche du Centre Hospitalier Universitaire de Sherbrooke, Sherbrooke, Québec, Canada; Faculté de génie, Université de Sherbrooke, Sherbrooke, Québec, Canada; Department of Critical Care Medicine, Sunnybrook Health Sciences Centre, Toronto, Canada; Interdepartmental Division of Critical Care Medicine, University of Toronto, Toronto, Canada; Faculté des sciences, Université de Sherbrooke, Sherbrooke, Québec, Canada; Department of Biostatistics, Epidemiology, and Informatics, Perelman School of Medicine, University of Pennsylvania, Philadelphia, PA, USA; Department of Medicine, Centre Hospitalier de l’Université de Montréal (CHUM), Montréal, Québec, Canada

**Author notes:** Corresponding author:* François Lamontagne, MD, MSc. Université de Sherbrooke, 3001 12e avenue Nord Sherbrooke, Québec, Canada.

**Keywords:** Arterial pressure targets, critical care, hypotension, vasopressor therapy

## Abstract

**Rationale:** The blood pressure value below which the benefits of vasopressors clearly outweigh their disadvantages is uncertain.

**Objectives:** The main objective of this analysis was to investigate the statistical properties and potential utility of indices estimating the vasopressor dose-rates as a function of blood pressure values over time.

**Methods:** In this single-center observational study, we collected blood pressure values from intensive care unit (ICU) monitors and norepinephrine dose-rates from infusion pumps corresponding to a derivation and a validation cohort. Patients included in each cohort were 18 years or older and received norepinephrine in the ICU. We defined and derived indices corresponding to vasopressor therapy above (>65 mmHg) and below (<60 mmHg) targets. We report the distribution of both indices over time from both cohorts as well as their associations with hospital mortality using logistic regression models adjusted for baseline variables.

**Results:** Between July 30 2020 and March 28 2022, 283 patients were included in the derivation cohort. The median ICU stay was 5 days (interquartile range [IQR] 2-10.5) and the median duration of norepinephrine therapy was 27.3 hours (IQR 13.9-54.2). The median cumulative time with a MAP below 60 mmHg was 0.9 hours (IQR 0.2-3.1), whereas the cumulative time with a MAP above 65 mmHg was 22.5 hours (IQR 10.9-40.4). Adjusting for prespecified baseline variables, the above target index was associated with hospital mortality (OR 1.6, 95% CI 1.2 –2.3) but not the below target index (OR 1.1 95% CI 0.8 – 1.5). Between May 5 2022 and December 16 2022, 83 patients were included in the validation cohort. The duration of ICU stay and the proportion of norepinephrine exposure above MAP target were consistent with the derivation cohort. The point estimates of the associations between each index and mortality were also consistent with the derivation cohort, but not statistically significant.

**Conclusion:** Indices of vasopressor administration as a function of blood pressure suggest that during most of vasopressor therapy episodes, patients are treated above target. The association of these indices with patient outcomes remains uncertain but could be explored in larger datasets, and future clinical trials of target MAPs may consider measurement of these indices to adjust results for treatment adherence.

## Introduction

The benefits of treatments aimed at correcting abnormal physiology are sometimes outweighed by their adverse effects. Antiarrhythmics for ectopic ventricular contractions following myocardial infarction,^1^ perioperative beta-blockers,^2^ blood transfusion thresholds above a hemoglobin value of 70 g/L,^3–6^ and the use of starch-based intravenous fluids^7,8^ constitute compelling examples of interventions that were, until recently, frequently administered to patients in intensive care units (ICUs) with the objective to restore ‘normal physiology’, until ultimately proven to be harmful.

The blood pressure value below which the benefits of vasopressors clearly outweigh their disadvantages is uncertain. Moreover, it is also unclear whether this threshold varies across subgroups. The value of vasopressor therapy in distributive shock is especially questionable because unless patients also suffer from hypovolemia, which may be easily corrected, vasodilation increases rather than decreases downstream perfusion.^9,10^ Clinical trials have thus far not confirmed the hypothesis that more aggressive vasopressor therapy improves outcomes in sepsis and other distributive states.^10–14^ Therefore, default practice should be to minimize vasopressor use. However, although a meta-analysis suggests that liberal vasopressor dosing regimens are not superior and could be inferior to more restrictive protocols,^14^ limitations related to differences in trial design and to imperfect protocol adherence complicate the interpretation of the results. In turn, observational data, which are often used to investigate the risks associated with vasopressor therapy and hypotensive episodes, are subject to residual confounding. Patients who are sicker are at the same time more likely to receive more vasopressors and to be hypotensive. A superior approach would consist of measuring the association with clinical outcomes of a variable that better reflects clinical decisions, while accounting for illness severity. We hypothesized that a variable combining vasopressor dose-rates and blood pressure values above and below prespecified thresholds would represent clinical decisions to achieve specific blood pressure values via pharmacological interventions.

The main objective of this analysis was to investigate the statistical properties and potential utility of indices that combine the dose of a vasopressor and the difference between a patient’s blood pressure and a prespecified target at any given point in time.

## Methods

### Populations

We studied two cohorts of patients. In both cohorts, patients were 18 years or older, treated at the medical intensive care unit (ICU) of the Centre intégré de santé et de services sociaux (CIUSSS) de l’Estrie – CHUS, and received intravenous norepinephrine (alone or in combination with other vasopressors) via a BBraun Infusomat infusion pump with DoseTrac software upgraded for this project (version 2.0). We limited this analysis to norepinephrine because it is the default vasopressor included in standard admission orders of the study ICU.

Patients in the <u>derivation cohort</u> were treated between July 30 2020 and March 28 2022 and could only be included if their blood pressure values were captured during vasopressor therapy, in part or in totality, via an arterial line. Patients in the <u>validation cohort</u> were treated between May 5 2022 and December 16 2022.

### Exposure – MAP-vasopressor indices

Standard admission orders in the study ICU specify that MAP values must be maintained between 60 and 65 mmHg, in keeping with recently published trials,^13^ meta-analyses,^14^ and guidelines.^15^ Clinicians were free to modify these orders for specific patients, for example, for patients prescribed supranormal MAP in the context of neurological injury. MAP values above 65 mmHg were defined as being ‘above target MAP’ whereas values below 60 mmHg were defined as ‘below target MAP’. Conceptually, we posited that any time vasopressors are administered while MAP values exceed 65 mmHg, a fraction (up to 100%) of the vasopressor dose-rate is administered in excess of the minimally effective dose to achieve the prespecified target. Since the precise fraction that is in excess is unknowable, the ‘above MAP-vasopressor index’ combines total norepinephrine dose-rates, MAP values greater than 65 mmHg, and time. To avoid misattributing to excessive vasopressor therapy rising MAP values after the resolution of the hypotensive episode, the index was only calculated during vasopressor infusions. Using this definition, the ‘above MAP-vasopressor index’ has a value greater than zero only when patients are receiving norepinephrine with MAP above 65 mmHg. Conversely, the ‘below MAP-vasopressor index’ combines MAP values below 60 mmHg, 1/vasopressor dose-rate, and time. To avoid misattributing low MAP values following the intentional discontinuation of vasopressors in the context of changes in the goals of care, (e.g. withdrawal of life-sustaining therapies), the ‘below MAP-vasopressor index’ was also calculated only during vasopressor therapy. To mitigate the impact of extreme MAP values, we used a non-linear function for the blood pressure component of both indices (atan(mmHg)^1.6^*mcg/kg for the ‘above MAP-vasopressor index’ and atan(mmHg)^1.6^*1/(mcg/kg) for the ‘below MAP-vasopressor index’). The mathematical expressions for these indices are detailed in the Online Data Supplement.

### Data sources and data handling

Several data sources were used for this pilot project (Fig 2). At inception, we anticipated that the indices would prove informative only if they were calculated using high-resolution data – i.e., vasopressor doses and MAP values collected every 5 seconds. However, because these high-resolution data were only available via a dedicated research infrastructure that was not readily available for all patients receiving vasopressors, we use various sources of lower-resolution data to mitigate the impact of missing high-resolution data.

#### High-resolution data

Paired data from ICU monitors (MAP) and infusion pumps (dose-rates derived from flow rates and norepinephrine concentrations) were collected in real-time via the Draeger Infinity HL7 high-speed and BBraun DoseTrac institutional server infrastructures. Data points were recorded approximately every 5 seconds and transferred in real-time to a proprietary data aggregation and synchronization software (custom Java application) and database (MariaDB developed by the MariaDB Foundation). In instances where norepinephrine dose-rates were recorded but at least one high-resolution MAP value was missing for no more than 60 minutes (e.g., due to disconnected arterial line), we imputed missing data points at 5-second intervals using linear interpolation between the closest available recorded values. In some instances, high-frequency data were missing for more than 60 minutes (e.g., because upgraded infusion pumps were not available, or no arterial lines were in place). We then resorted to various sources of lower frequency data.

#### Missing high-resolution data in the derivation phase

Hourly MAP and norepinephrine dose-rate values that were charted by ICU nurses were retrieved from the patients’ medical records for missing segments of norepinephrine therapy of 4 hours or more at the beginning or the end of vasopressor episodes.

#### Missing high-resolution data in the validation phase

During the validation phase of the study, a second source of low-resolution data consisted of MAP values that were measured via non-invasive blood pressure cuffs (as opposed to arterial lines) directly from the ICU monitors. These MAP values were used when arterial line values were missing and the gap between available MAP values (either from arterial line of non-invasive cuff) was no more than 60 minutes.

In contrast to the derivation phase, the nurse-charted hourly MAP and norepinephrine dose-rate data were collected for the entire duration of norepinephrine episodes. Therefore, for any interval of >1 hour of missing high-resolution data, linear interpolation was used to fill in missing values using the closest available high-resolution and manually charted data points.

#### Clinical data

Two members of the research team independently reviewed medical records to collect all-cause mortality at hospital discharge (primary outcome) and the following variables at ICU admission: age, sex, APACHE II score, admission diagnosis, and chronic comorbidities (hypertension, atherosclerosis, heart failure and chronic dialysis).

### Statistical analyses

#### Descriptive analyses

We describe episodes of norepinephrine infusions, specifically dose-rates, MAP values, and corresponding ‘above and below target indices’ over time. We report continuous variables as means (standard deviations) or medians (first and third quartiles), as appropriate, and categorical variables as counts (percentages). The distribution of patient-level ‘above and below target indices’ vasopressor exposure indices over time was plotted separately for decedents and survivors to facilitate visual comparisons.

#### Correlation between MAP-vasopressor indices derived from high and low automated sampling frequency

In both cohorts, we evaluated the correlation between MAP-vasopressor indices calculated using high and low-frequency sampling of paired MAP and norepinephrine values. To do this, we restricted our analysis of the high-resolution dataset to values collected at one-hour intervals (i.e., first low-resolution dataset consisted of the first pair of MAP and vasopressor dose-rates at the beginning of each hour and ignoring the subsequent data pairs collected every 5 seconds until the following hour). We then estimated above and below target indices using data from the resulting high and low-frequency sampling frame and estimated their correlation using Kendall’s tau.

#### Correlation between indices derived from automated data sampling vs charted data

During the validation phase of the study, we evaluated the correlation between MAP-vasopressor indices calculated using low-frequency sampling of paired MAP and norepinephrine values collected directly from ICU monitors and infusion pumps (a combination of first and second low-resolution dataset) and all low-frequency data charted by ICU nurses and retrieved manually from the medical records (third low-resolution dataset). For automated sampling, we calculated MAP-vasopressor indices using hourly values of norepinephrine dose-rates from infusion pumps and hourly MAP values from the high-resolution dataset or, when these were not available, MAP values measured via non-invasive blood pressure cuffs collected directly from ICU monitors. For manual sampling, we calculated vasopressor indices using data charted by ICU nurses and retrieved manually from medical records. The correlation between MAP-vasopressor indices from these two data sources was measured using Kendall’s tau.

#### Exploring associations between MAP-vasopressor indices and all-cause in-hospital mortality

For each of the derivation and validation cohorts, we conducted two multivariable logistic regression models with in-hospital mortality as the dependent variable. One model used the ‘above target MAP-vasopressor index’ as the main exposure, and the second, the ‘below target MAP-vasopressor index’. In both models, the association between indices and mortality were adjusted for the same prespecified baseline clinical characteristics: age, APACHE II, hypertension, chronic dialysis, atherosclerotic disease, sex and heart failure. For these analyses, all continuous variables were centered and scaled.

R version 4.3.1 (R Foundation for Statistical Computing, Vienna, Austria) was used for the statistical analyses.

### Ethics

The Research Ethics Board of the CIUSSS-Estrie reviewed the protocol of this cohort study and waived the need to obtain informed consent to prospectively collect high resolution anonymized data on MAP and vasopressor dose-rates on the condition that linkage with data from the patients’ medical records (e.g., age, sex, comorbidities, outcomes) occur retrospectively (i.e., after hospital discharge).

## Results

### Derivation cohort

From July 30 2020 to March 28 2022, 1722 patients were hospitalized in the participating intensive care unit. Of those, 686 were prescribed norepinephrine (40%) and 316 (46% of patients prescribed norepinephrine) were treated with one of the infusion pumps upgraded for this study. After excluding patients whose data could not be retrieved (e.g. no arterial line, upgraded infusion pumps unavailable) or who were organ donors, data from 283 patients (41% of patients prescribed norepinephrine) contributed to this analysis. The inclusion flow diagram is depicted in Figure 1.

**Figure 1.**
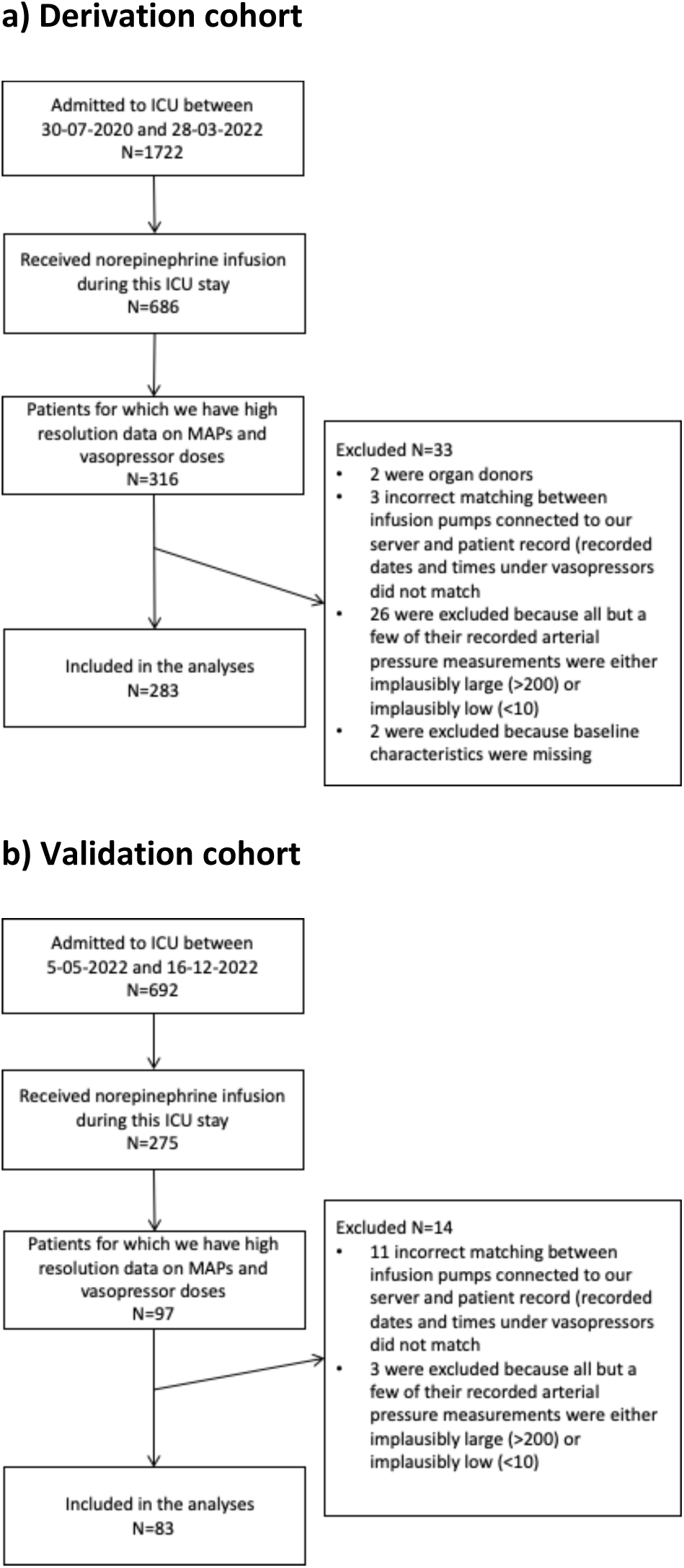
Flow diagram.

**Figure 2.**
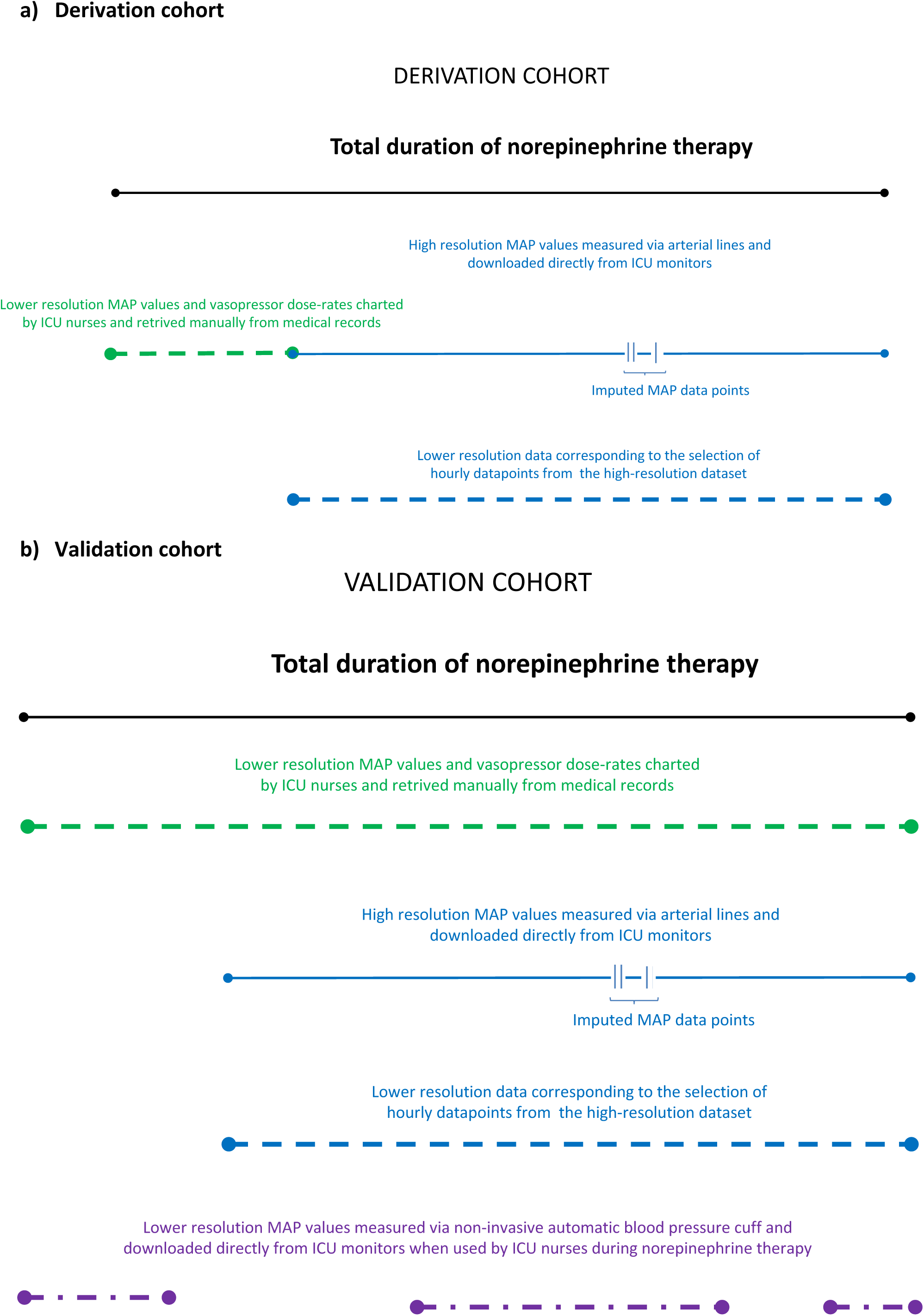
Data sources. <u>High-resolution data</u>: Paired data from ICU monitors (MAP) and infusion pumps (dose-rates derived from flow rates and norepinephrine concentrations) were collected in real-time via the Draeger Infinity HL7 high-speed and BBraun DoseTrac institutional server infrastructures. Data points were recorded approximately every 5 seconds. In instances where norepinephrine dose-rates were recorded but at least one high-resolution MAP value was missing for no more than 60 minutes (e.g., due to disconnected arterial line), we imputed missing data points at 5-second intervals using linear interpolation between the closest available recorded values. In some instances, high-frequency data were missing for more than 60 minutes (e.g., because upgraded infusion pumps were not available, or no arterial lines were in place). We then resorted to various sources of lower frequency data. <u>Low-resolution MAP values and vasopressor dose-rates charted by nurses</u>: Hourly MAP and norepinephrine dose-rate values that were charted by ICU nurses were retrieved from the patients’ medical records. In the derivation cohort, this was done for missing segments of norepinephrine therapy of 4 hours or more. In the validation cohort, this was collected for the entire duration of the vasopressor episode. <u>Low-resolution MAP values measured automatically via non-invasive blood pressure cuffs</u>: In the validation cohort, when arterial line values were missing MAP values measured via non-invasive blood pressure cuffs were downloaded directly from the ICU monitors. <u>Low-resolution selection of data in the high-resolution dataset</u>: To investigate the correlation between indices measured using high- vs. low-resolution sampling frames, we created duplicates of both high-resolution datasets that were restricted to hourly datapoints.

Baseline characteristics are reported in Table 1. The most common admission diagnosis was sepsis, followed by cardiovascular, and respiratory disorders (Table 1). The median length of stay in ICU was 5 days (interquartile range [IQR] 2, 10.5), and 114 patients (40.3%) died in hospital. The median duration of norepinephrine therapy was 27.3 hours (IQR 13.9 – 54.2). Table 2 summarizes the median durations of norepinephrine episodes captured by high- and low-resolution sampling frequency.

**Table 1.**
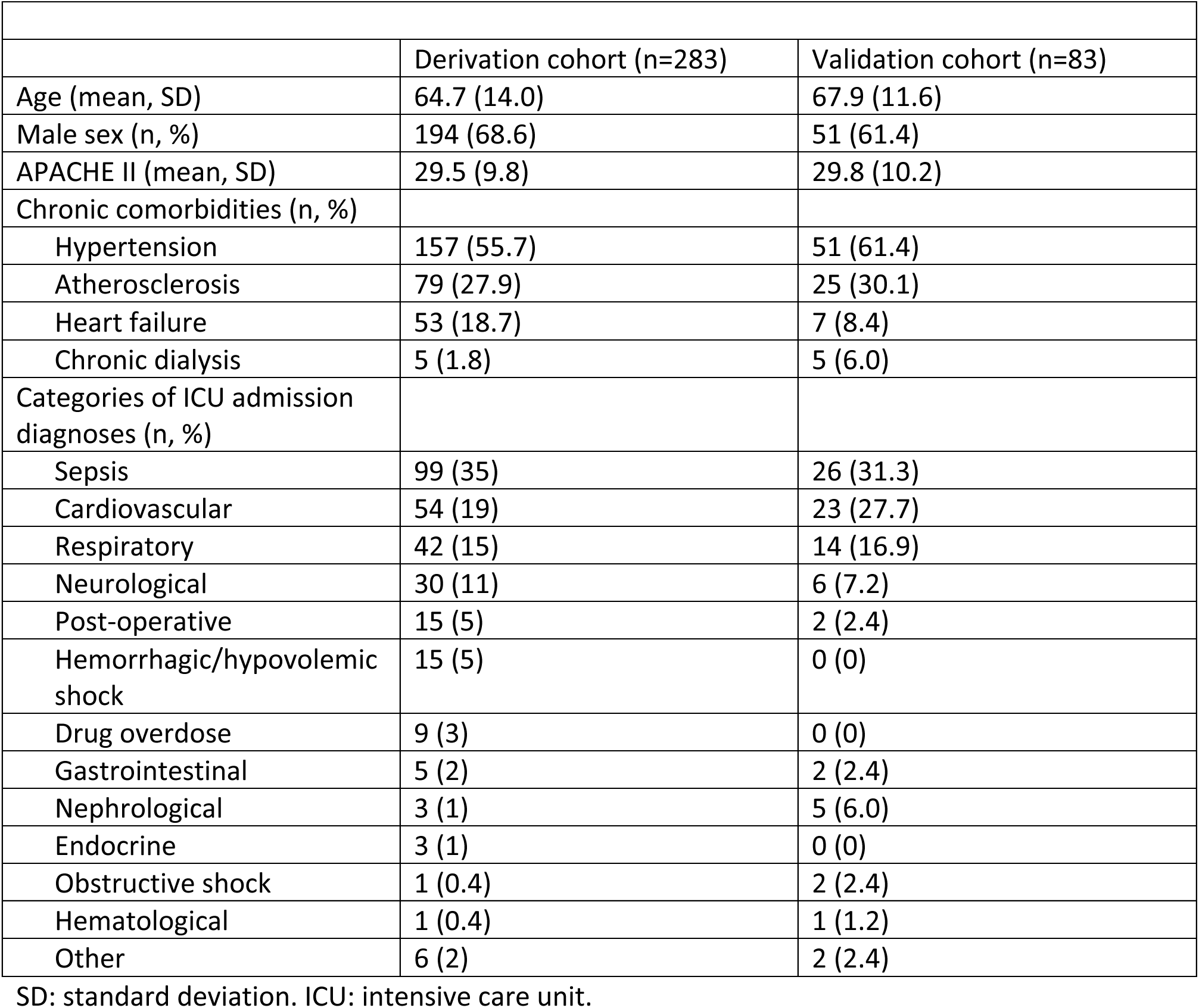
Baseline characteristics of patients enrolled the derivation and validation cohorts.

**Table 2.** Duration of norepinephrine therapy and data sources in the derivation and validation cohorts.

|  | Derivation cohort (n=283) | Validation cohort (n=83) |
| --- | --- | --- |
| Total duration of norepinephrine therapy (hours); median, IQR | 27.3 (13.9 – 54.2) | 34.5<sup>1</sup> (13.3 – 55.8) |
| Duration of high-resolution data capture (hours); median, IQR | 18.4<sup>1</sup> (9.1 – 39.3); n=283 | 16.7<sup>2</sup> (3.4 – 41.1); n=83 |
| Duration of low-resolution data capture based on MAP and norepinephrine dose-rate values charted hourly by ICU nurses (hours); median, IQR | 11.0 (7.0 – 23.0); n=157 | 34.0<sup>2</sup> (16.3 – 53.6); n=83 |
| Proportion of total duration of norepinephrine therapy captured by high-resolution data (%); median, IQR | Among all patients: 90.1 (55.3 – 100.0); n=283<br>Including patients that had data retrieved using nursing charts: 59.3 (36.0 – 76.5); n=157 | Among all patients: 67.8 (33.2 – 97.7); n=83 |

<sup>1</sup> Linear interpolation was used to impute missing MAP datapoints in 274 patients (96.8% of the derivation cohort) to impute a median of 12 missing high-resolution MAP values (0.5% of total high-resolution MAP values) per patient (IQR 5 – 22). When imputation was applied, the median of the patient-average time interval separating available high-resolution data points from either side of a missing value sequence was of 26.4 seconds (IQR 18.1 – 37.8).
<sup>2</sup> Linear interpolation was used to impute missing MAP datapoints in 48 patients (58% of the validation cohort) to impute a median of 14.5 missing high-resolution MAP values (0.5% of total high-resolution MAP values) per patient (IQR 7.0 – 33.0). When imputation was applied, the median of the patient-average time interval separating available high-resolution data points from either side of a missing value sequence was of 26.7 seconds (IQR 19.2 – 40.1).
<sup>3</sup> In the validation cohort, the entire duration of norepinephrine episodes was captured by retrieving hourly data charted by ICU nurses; the difference between the total duration of norepinephrine therapy and the duration of the low-resolution data capture is explained by the loss of precision attributable to the hourly data capture. IQR: interquartile range

#### MAP values during vasopressor therapy

The mean patient-average MAP during vasopressor therapy was 73.2 mmHg (SD 6.7) (Table 3). On average, across patients, MAP values were below 60 mmHg for 6.2% (SD 10.8%) of the total duration of the norepinephrine episode, while they were above 65 mmHg for 80.9% (SD 17.1%) and in-range for 12.9% (SD 10.6%) of the episode (Figure 3).

**Figure 3.**
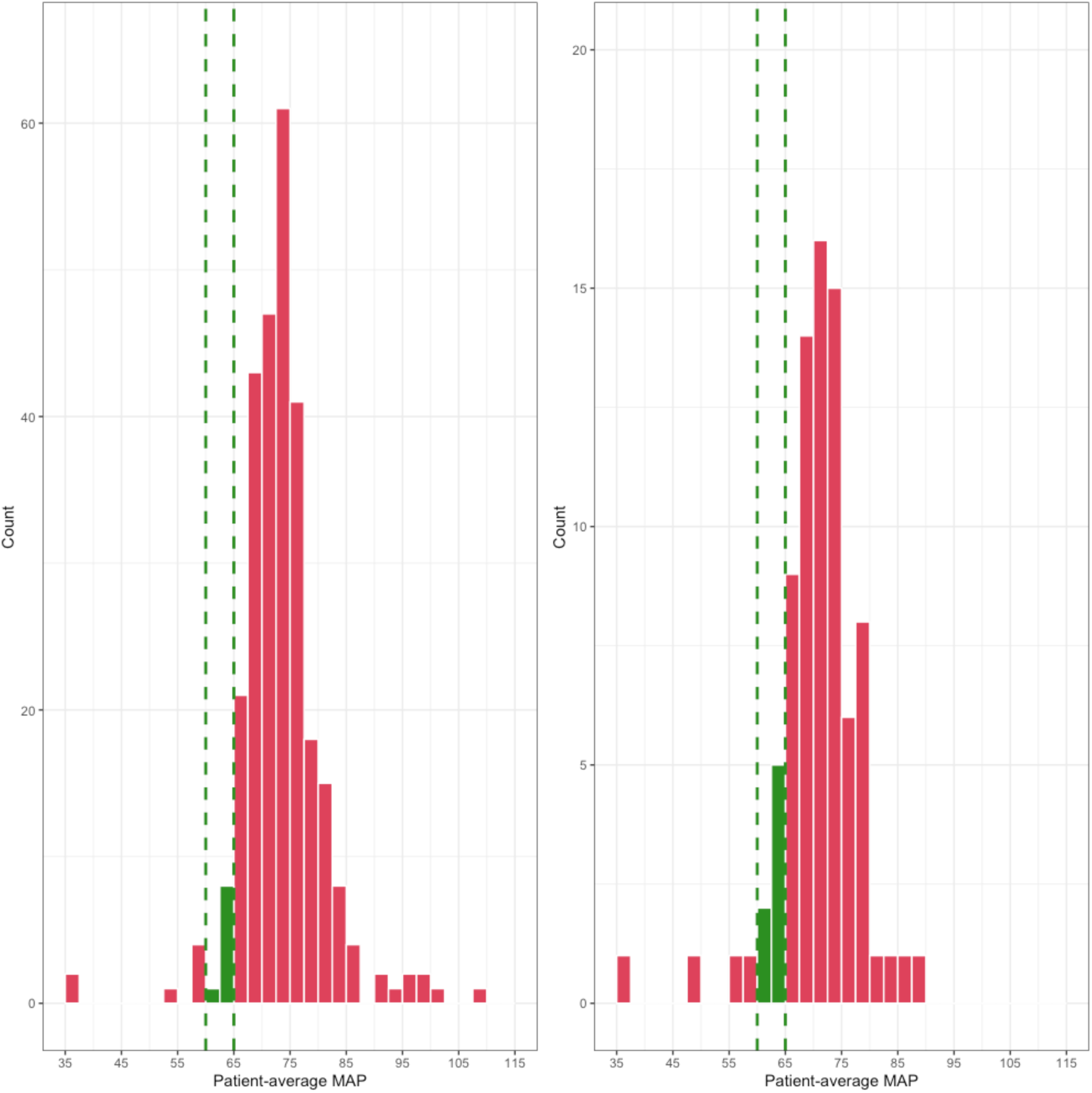
Distribution of MAP values (Derivation cohort: n=283 patients; validation cohort: n=83) Histograms depicting the distribution of patient-average MAP values during vasopressor therapy. Left panel: derivation cohort. Right panel: validation cohort.

**Table 3.** MAP values during vasopressor therapy.

|  | Derivation cohort<sup>1</sup> (n=283) | Validation cohort<sup>2</sup> (n=83) |
| --- | --- | --- |
| Patient-average MAP during norepinephrine therapy (mmHg); mean, SD | 73.2 (6.7) | 72.1 (7.0) |
| Cumulative patient duration of norepinephrine therapy episodes with MAP values below 60 mmHg (hours); median, IQR | 0.9 (0.2 – 3.1) | 1.3 (0.3 – 4.2) |
| Percentage of total norepinephrine therapy duration of episodes with MAP values below 60 mmHg (%); median, IQR | 2.8 (1.0 – 7.1) | 5.0 (1.1 – 11.3) |
| Cumulative patient duration of norepinephrine therapy episodes with MAP values between 60 and 65 mmHg (hours); median, IQR | 2.7 (0.9 – 7.6) | 3.8 (1.1 – 9.1) |
| Percentage of total norepinephrine therapy duration of episodes with MAP values between 60 and 65 mmHg (%); median, IQR | 10.2 (4.7 – 18.5) | 12.4 (6.1 – 18.2) |
| Cumulative patient duration of norepinephrine therapy episodes with MAP values above 65 mmHg (hours); median, IQR | 22.5 (10.9 – 40.4) | 24.4 (12.9 – 43.5) |
| Percentage of total norepinephrine therapy duration of episodes with MAP values above 65 mmHg (%); median, IQR | 86.7 (72.6 – 92.1) | 82.6 (70.8 – 91.8) |

<sup>1</sup> Estimates were calculated using values in the high-frequency sampling dataset when available and from hourly MAP values charted by ICU nurses and retrieved from medical records when high-frequency data were not available. <sup>2</sup> Estimates were calculated using values in the high-frequency sampling dataset when available and from low-frequency MAP values measured via non-invasive blood pressure cuffs collected directly from ICU monitors nurses and retrieved from medical records when high-frequency MAP data were not available. During segments of norepinephrine infusion that were not captured by upgraded pumps, we retrieved norepinephrine dose-rates charted by ICU nurses from the medical records.

#### Norepinephrine dose-rates and MAP-vasopressor indices

The median patient-average norepinephrine dose-rate was 5.8 μg/min (IQR 2.9–10.2) and the median cumulative dose of norepinephrine received was 10879.8 μg (IQR 3315.3–26270.3). Figure 4 illustrates the distribution of the above and below target vasopressor indices. Figure 5 illustrates distribution of the duration of time in each quartile of above and below MAP-vasopressor index for each patient, by vital status at hospital discharge.

**Figure 4.**
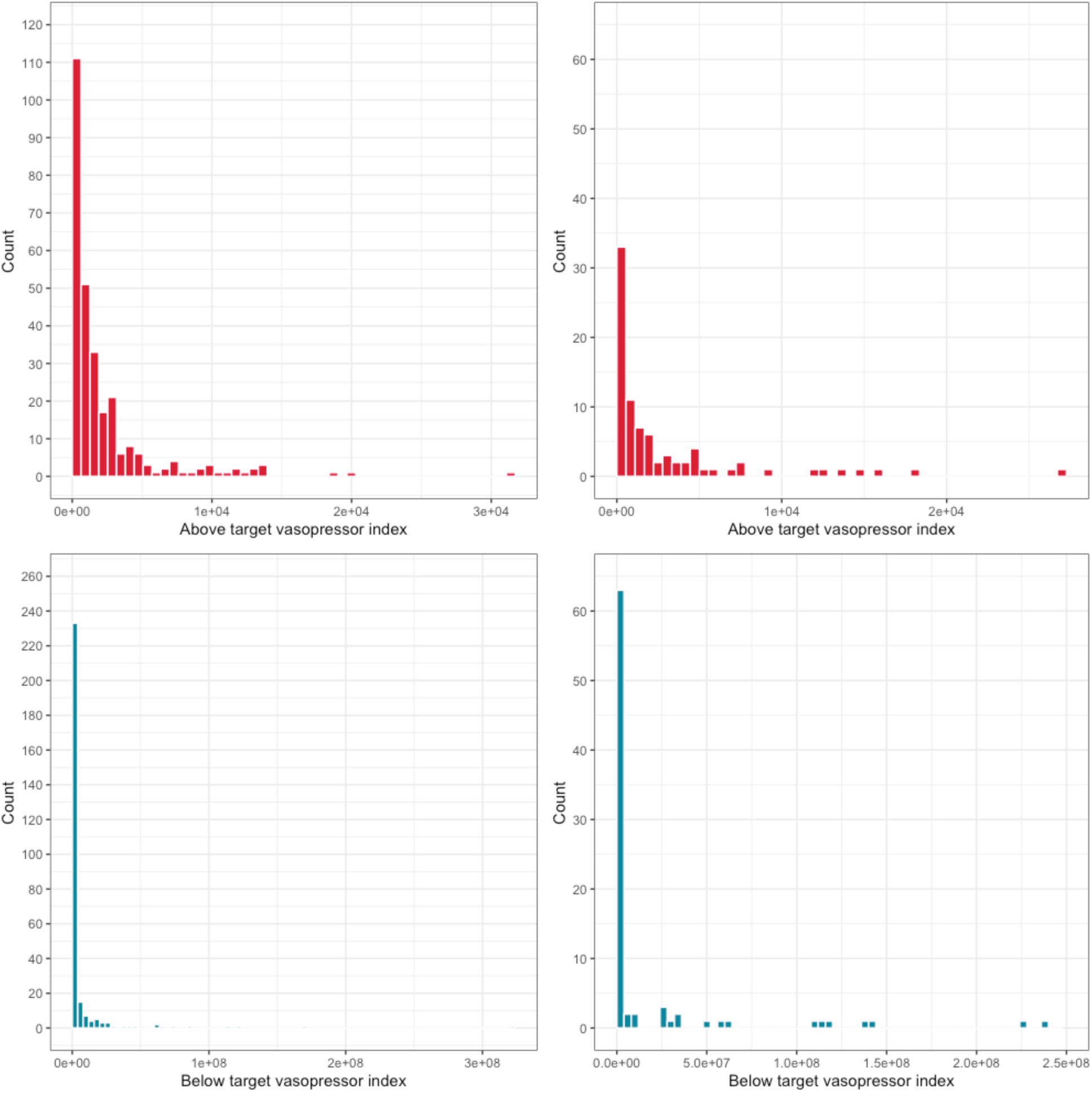
Distribution of vasopressor therapy indices (Derivation cohort: n=283 patients; validation cohort: n=83) Histograms depicting the distribution of the vasopressor therapy indices. Upper panel: ‘above 65’ index in the derivation cohort (left) and in the validation cohort (right). Bottom panel: ‘below 60’ index in the derivation cohort (left) and in the validation cohort (right).

**Figure 5.**
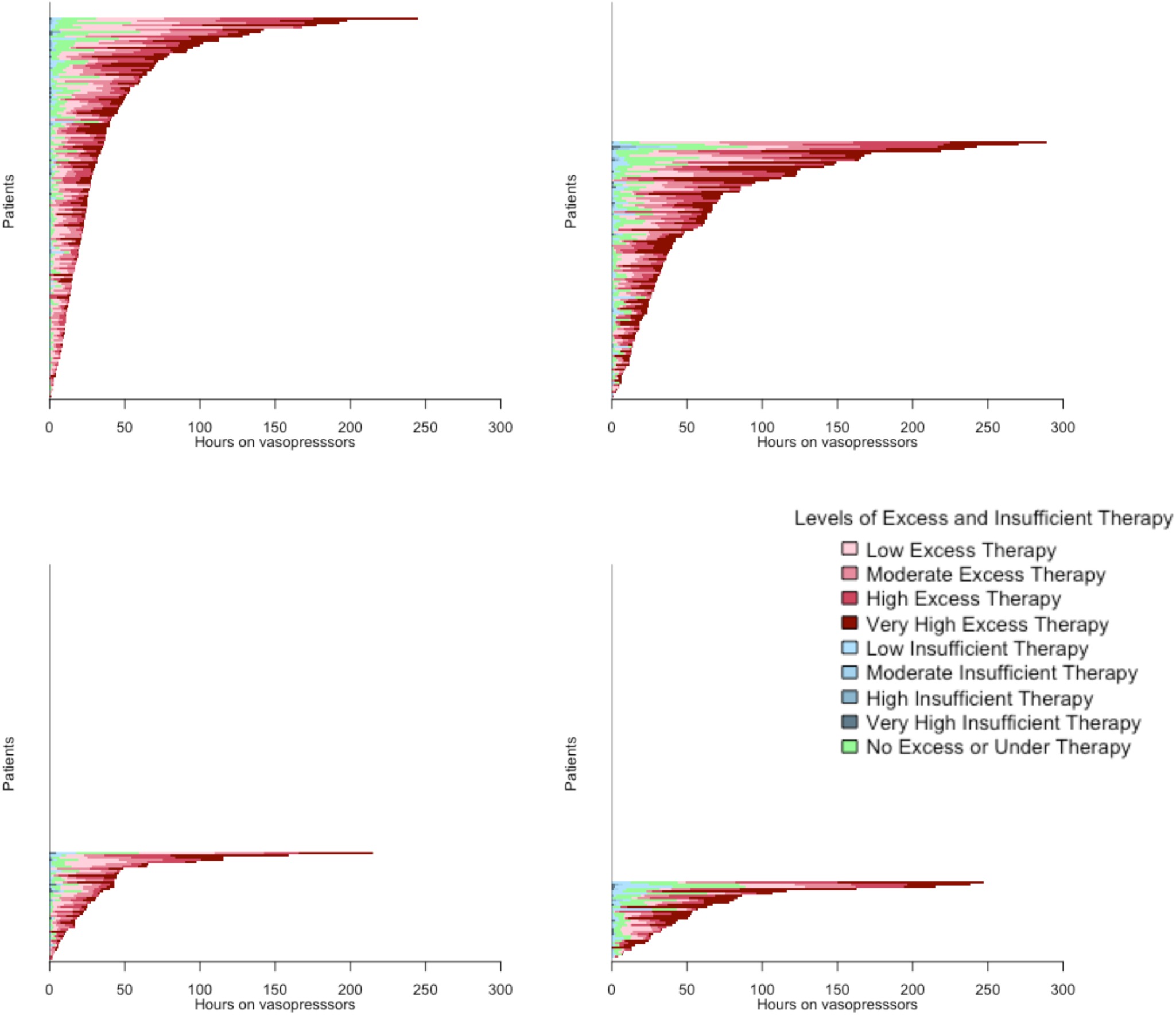
Distribution of time under vasopressor therapy relative to levels of ‘excess’ and ‘insufficient’ therapy (Derivation cohort: n=283 patients, 169 survivors, 114 deceased; validation cohort:n=83, 48 survivors, 35 deceased) Each line represents a patient, ordered by descending order of total duration of vasopressor therapy. For each patient, the duration of time in each quartile of above MAP-vasopressor index (of red) and below MAP-vasopressor index (of blue) is shown. The upper two panels display the derivation cohort and the lower two panel the validation cohort. Within each cohort, survivors are displayed on the left and decedents on the right. There is a higher density of dark red, indicating high to very high excess vasopressor therapy, in the panels for decedents compared to survivors.

**Alternative to Figure 5:**
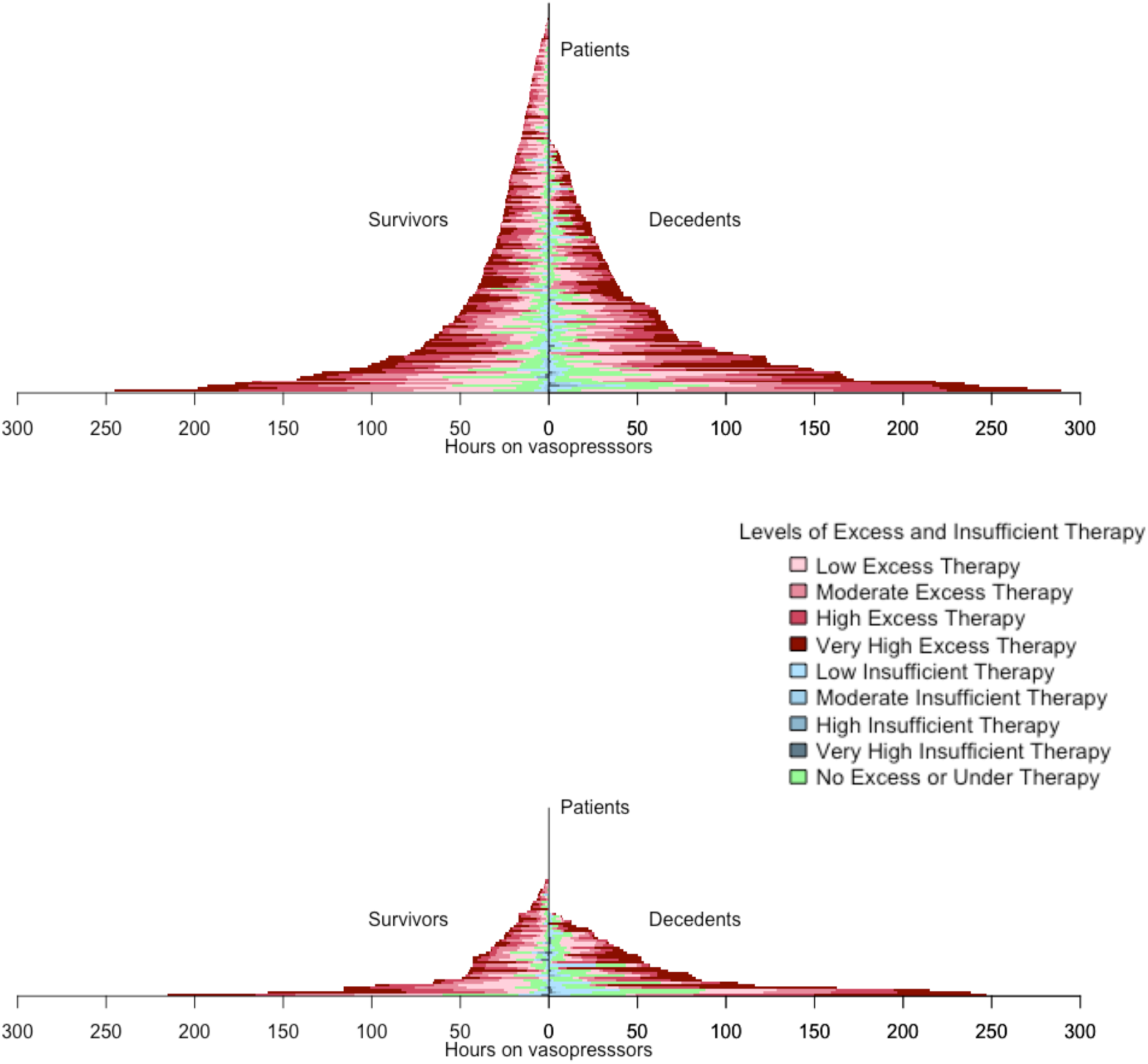
Each line represents a patient, ordered by ascending order of total duration of vasopressor therapy. For each patient, the duration of time in each quartile of above MAP-vasopressor index (of red) and below MAP-vasopressor index (of blue) is shown. The upper panel display the derivation cohort and the lower panel the validation cohort. Within each cohort, survivors are displayed on the left and decedents on the right. There is a higher density of dark red, indicating high to very high excess vasopressor therapy, in the panels for decedents compared to survivors.

The correlation between high and low resolution estimates of above and below MAP-vasopressor indices was 0.79 (95% CI 0.75 – 0.82) and 0.52 (95% CI 0.47 – 0.58), respectively.

#### Associations between MAP-vasopressor indices and mortality

Adjusting for age, APACHE II, hypertension, chronic dialysis, atherosclerotic disease, sex and heart disease/heart failure, the above MAP-vasopressor index was associated with hospital mortality (OR 1.6 for each SD, 95% CI 1.2 –2.3) but not the below target index (OR 1.1 95% CI 0.8 – 1.5) (Table 4).

**Table 4.** Association between above and below MAP-vasopressor indices and hospital mortality.

|  | Derivation cohort | Validation cohort |
| --- | --- | --- |
|  | OR (95% CI) | OR (95% CI) |
| Above MAP-vasopressor index<sup>1</sup> | 1.6 (1.2 – 2.3) | 1.7 (0.9 – 4.0) |
| Below MAP-vasopressor index<sup>1</sup> | 1.1 (0.8 – 1.5) | 1.4 (0.7–2.6) |

<sup>1</sup> Adjusted for Age, APACHE II, Hypertension, Chronic dialysis, Atherosclerosis, Heart disease/heart failure, and Sex (see Table E.1 in the online supplement for the value of the adjusting variables fitted coefficients.)

### Validation cohort

From May 5 2022 to December 16 2022, 692 patients were treated in the ICU. Of those, 275 received norepinephrine (40%) and 97 (35% of patients prescribed norepinephrine) were treated with one of the infusion pumps upgraded for this study. After excluding patients whose data could not be retrieved (e.g. limited access to research pumps) or who were organ donors, data from 83 patients (30% of patients who received norepinephrine between May and December 2022) contributed to this analysis (Figure 1b). The most common admission diagnosis was sepsis, followed by cardiovascular, and respiratory disorders. The median length of stay in ICU was 5 days (IQR 2.5, 8.0), and 35 patients (42.2%) died in hospital. The median duration of norepinephrine therapy was 34.5 hours (IQR, 13.3, 55.8) (Table 2).

#### MAP values during vasopressor therapy

The mean patient-average MAP during vasopressor therapy was 70.9 mmHg (SD 7.2) (Table 3). On average, across patients, MAP values were below 60 mmHg for 9.2% (SD 14.4%) of the total duration of the norepinephrine episode, while they were above 65 mmHg for 76.9% (SD 20.8%) of the time, and in-range for 13.9% (SD 11.1%) of the time (Figure 3).

#### Norepinephrine dose-rates and MAP-vasopressor indices

The median patient-average norepinephrine dose-rate was 6.2 μg/min (IQR 3.6 – 10.8). Figure 4 illustrates the distributions of the patient-average cumulative MAP-vasopressor indices and Figure 5 illustrates the duration of indices for each patient overall and then by survival at hospital discharge.

The correlation between high and low resolution estimates of above and below MAP-vasopressor indices was 0.74 (95% CI 0.64 – 0.84) and 0.50 (95% CI 0.39 – 0.61), respectively.

#### Associations between vasopressor indices and mortality

Adjusting for age, APACHE II, hypertension, chronic dialysis, atherosclerotic disease, sex, and heart failure, point estimates for the above MAP-vasopressor index (OR 1.7, 95% CI 0.9 – 4.0) and the below MAP-vasopressor index (OR 1.4, 95% CI 0.7–2.6) were consistent with those in the derivation cohort, but not statistically significant (Table 4).

## Discussion

In this analysis of real-world data from critically ill patients treated with norepinephrine, we found that MAP values were maintained above the prescribed blood pressure targets for 80 to 90% of the total time on vasopressors. Because both indices estimate the effects of vasopressors as a function of blood pressure values, they may be better suited to investigate optimal blood pressure targets than MAP values or vasopressor doses alone, which are both subject to significant residual confounding.

Our results are consistent with other reports describing MAP values during vasopressor therapy, which have shown that clinicians seldom allow MAP values to drop below 70 mmHg, even if this entails administering higher doses of catecholamines for longer periods.^16–18^ In the most recent trial of MAP targets in critically ill patients, the average MAP observed in the usual care control arm (1307 patients) treated at the discretion of ICU teams was 73 mmHg whilst receiving vasopressors.^13^ The indices we describe represent a direct measure of vasopressor treatment intensity, which is a modifiable clinical variable. Measures of associations between the proposed indices and mortality are exploratory, but they too are consistent with effect estimates of clinical trials comparing the higher vs. lower blood pressure targets for vasopressors.^14^

Strengths of this analysis include the use of novel indices accounting for both aspects of vasopressor therapy: dose and corresponding blood pressure values, fill a gap in cardiovascular resuscitation research. Another strength lies in the applicability of these measures to observational as well as clinical trial datasets. In the context of observational studies, the indices may be used to investigate the effects of vasopressors as a function of blood pressure and the optimal blood pressure target in various subgroups. In clinical trials, they may constitute measures of protocol adherence.

We also note the following limitations. We collected data from two small cohorts of patients treated at the ICU of a single center, which limits the generalizability of our results but was appropriate given the pilot and exploratory nature or our work. We acknowledge that we could not obtain data on a large proportion of all eligible patients and that we were forced to impute missing data among the patients that were included in this analysis. This is largely due to the limited availability of the infrastructure required for high-resolution data-capture. Correlations between high- and low-resolution datasets in this study allowed us to quantify the weight of information that is lost when hourly data points are used rather than a higher frequency sampling frame. While higher-resolution datasets may be necessary for some purposes (e.g., for machine learning models and real-time automated closed-loop vasopressor delivery systems), hourly data may be sufficient to characterize adherence to trial protocols and to monitor usual care practices and their effects in large observational cohorts. The vasopressor indices developed assume that the individual factors of difference of MAP from target, norepinephrine dose-rate, and time contribute equally to the effect, but alternative data science approaches in larger datasets may find that a different weighting leads to a higher association with mortality. As mentioned, associations between reported indices and mortality are, for the moment, strictly exploratory. In particular, we note that our analysis provides almost no information on the effects of reducing vasopressor therapy intensity (i.e., ‘below target’ index) as clinicians almost never allowed the patients’ MAP to drop below 60 mmHg.

In conclusion, Indices of vasopressor administration as a function of blood pressure suggest that during most of vasopressor therapy episodes, patients are treated above target. The association of these indices with patient outcomes remains uncertain but could be explored in larger datasets, and future clinical trials of target MAPs may consider measurement of these indices to adjust results for treatment adherence.

## Supporting information

Highres Supplement

## Data Availability

All data produced in the present study are available upon reasonable request and submission of detailed analysis plan to the authors

Alain Gervais: conception, design, data acquisition, writing - original draft;

François Lamontagne: conception, design, data acquisition, data analysis, data interpretation, writing - original draft;

Jean-Baptiste Michaud: conception, design, data acquisition, writing - review and editing;

Neill KJ Adhikari: data interpretation, writing - original draft, writing - review and editing;

Jean-Michel Pagé: data analysis, writing - review and editing;

Marie-Hélène Masse: data acquisition, writing - review and editing;

Michael O Harhay: data interpretation, writing - review and editing;

Michaël Chassé: data interpretation, writing - review and editing;

Félix Lamontagne: data acquisition, writing - review and editing;

Katia Laforge: data acquisition, writing - review and editing;

Alexandra Fortin: data acquisition, writing - review and editing;

Marc-André Leclair: data acquisition, writing - review and editing;

Simon Lévesque: data analysis, writing - review and editing;

Marie-Pier Domingue: data analysis, writing - review and editing;

Neda Momenzadeh: data analysis, writing - review and editing;

Martin Vallières: data interpretation, writing - review and editing;

Ruxandra Pinto: data interpretation, writing - review and editing;

Maxime Morin-Lavoie: data interpretation, writing - review and editing;

Francis Carter: data acquisition, writing - review and editing;

Félix Camirand Lemyre: conception, design, data acquisition, data analysis, data interpretation, writing - original draft.

## Data Sharing Statement

Individual, de-identified patient data will not be shared, as we do not have the necessary permissions to do so.

This article has an online data supplement.

