## Supplementary material for "Derivation and validation of indices incorporating vasopressor dose and blood pressure values over time": Highres Supplement

### Reliability assessments for clinical data collection

For every binary variable considered in this analysis, we computed Cohen's kappa to evaluate the level of agreement between raters, and a value of 0.8 or higher was achieved for most of them, except for admission diagnosis and heart disease. For these variables, an additional member of the research team independently reviewed the medical records to resolve the conflicting outcomes.

Regarding continuous variables, we determined the proportion of matching values in the recorded data. All presented a proportion above 0.8 and were thus deemed reliable.

### 'Excess' and 'insufficient' vasopressor therapy indices

The 'excess vasopressor therapy index' is a measure based on three factors: mean arterial pressure (MAP) above 65 mmHg, vasopressor dose-rate, and time. It is used to quantify the degree of excessive vasopressor therapy. This index only has a value greater than zero when the MAP is above 65 mmHg, indicating that the therapy may be excessive. The unit for this index is s

= atan(mmHg)$^{1.6}$*mcg/kg, which is a mathematical function combining the MAP and the vasopressor dose-rate.

The 'insufficient vasopressor therapy index' is another measure that represents the product of MAP below 60 mmHg, the inverse of vasopressor dose-rate (1/vasopressor dose-rate), and time. The unit for this index is atan(mmHg)$^{1.6}$*1/(mcg/kg). This index is used to quantify the level of insufficient vasopressor therapy, which can occur when the MAP is below 60 mmHg.

To account for the impact of extreme MAP values on these indices, a non-linear function was used for the blood pressure component in both products. This helps provide a more accurate representation of the relationship between blood pressure and the therapy indices.

To ensure that the insufficient vasopressor therapy index accurately reflects the therapy levels and does not mistakenly include cases where vasopressors were intentionally discontinued (e.g., withdrawal of life-sustaining therapies), these indices were only calculated when patients were actively receiving vasopressor therapy.

**Mathematical derivation of the 'Excess' vasopressor index**

As mentioned earlier, for a given patient, the 'Excess' vasopressor index is calculated from MAP and dose rate measurements recorded over time. Let $t_i$ represent the timestamp of the i$^{th}$ measurement of a given patient, $T$ their associated total number of measurements, and $MAP_{t_i}$

and $DR_{t_i}$ their average arterial pressure and dose rate measured on the i$^{th}$ occasion (i.e., at time $t_i$ ).

Conceptually, a patient's 'Excess' vasopressor index (EVI) calculated over a given time interval $[t_i, t_{i+1}]$ is designed to capture the total amount of vasopressors received during the latter time window, weighted by a factor that takes the value 0 when their MAP is below the target (i.e., when the full dose is needed and is not in 'Excess') and proportional to how far their MAP is from the target otherwise (i.e., when a portion of the dose is in excess). A formula that would implement this idea would take the form

$$EVI_{[t_i,t_{i+1}]} = \max\left(MAP_{[t_i,t_{i+1}]} - 65{,}0\right) * DR_{[t_i,t_{i+1}]} * (t_{i+1} - t_i)$$

where $MAP_{[t_i,t_{i+1}]}$ and $DR_{[t_i,t_{i+1}]}$ respectively denote their average MAP and dose rate value over the time interval $[t_i, t_{i+1}]$, i.e.,

$$MAP_{[t_i,t_{i+1}]} = (MAP_{t_i} + MAP_{t_{i+1}})/2$$

$$DR_{[t_i,t_{i+1}]} = (DR_{t_i} + DR_{t_{i+1}})/2$$

and where the term $(t_{i+1} - t_i)$ represents the length of the time interval. A patient's 'Excess' vasopressor therapy index would then be obtained as the sum of all their interval-specific excess therapy values defined above, i.e.,

$$EVI = \sum_{i=1}^{T-1} EVI_{[t_i,t_{i+1}]} \qquad (1)$$

However, operating in this way would have meant attributing overly large weights to perfused doses for periods of time where the MAP of a patient would have been measured far from the actual target. To mitigate the impact of MAP measurements far from the target, we used a non-linear function that mimics the above formula when MAP values are near the target, but possesses an asymptote, which has the effect of capping the weight attributed to large MAP values. Formally,

$$EVI_{[t_i,t_{i+1}]} = w * \left( atan\left( max\left(MAP_{[t_i,t_{i+1}]} - 65{,}0\right) * \frac{\pi}{2w} \right) \right)^z * DR_{[t_i,t_{i+1}]} * (t_{i+1} - t_i)$$

*( 2)*

where $w$ and $z$ are constants chosen to make the resulting variable as close as possible to the one using only the weighting factor $\max\left(MAP_{[t_i,t_{i+1}]} - 65{,}0\right)$ when MAP values are in the range [65, 65+w]. The latter formula ensures the two versions of the Excess Therapy variable coincides at 65 and at 65+w, as well as for all MAP values below 65. The value of $w$ was set so as to make $65 + w$ equal to the $90^{th}$ percentile of the patient-average MAP values and we subsequently chose the value of $z$ that minimized the area under the curve of the squared difference between the function

$$x \rightarrow \; w * \left( atan\left( x * \frac{\pi}{2w} \right) \right)^z$$

*(3)*

and

$$x \rightarrow x$$

on [0, w], resulting in the values $w = 14$ and $z = 1.6$. The curve

$$MAP \; \rightarrow w * \left( atan\left( max(MAP - 65{,}0) * \frac{\pi}{2w} \right) \right)^z$$

computed with these values for $w$ and $z$ is illustrated below against the curve

$MAP \rightarrow \max(MAP - 65{,}0)$. Figure E1 illustrates the effect of the non-linear transformation used in the 'excess' therapy index.

**Figure E1. Graphical representation of the effect of the non-linear transformation used in the 'Excess' therapy index.**

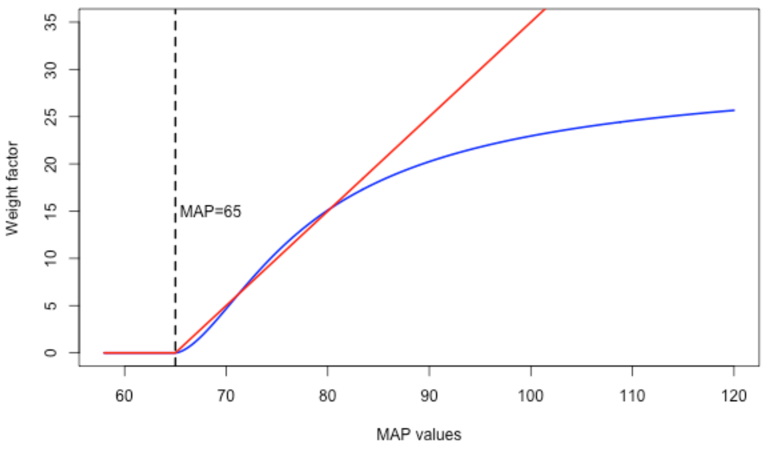

The blue curve represents the non-linear transformed values of the term $max(MAP - 65{,}0)$ following the formula in equation (3). The red line represents the curve that is linear in the term $max(MAP - 65{,}0)$ .

From there, a patient's total excess therapy is calculated using Equation (1), with $EVI_{[t_i,t_{i+1}]}$ as in Equation (2).

**Mathematical derivation of the 'Insufficient' vasopressor therapy index**

The 'insufficient' vasopressor index (IVI) is design to mimic the EVI, except for the dose rate term. Formally, the IVI of a given patient on the time interval $[t_i, t_{i+1}]$ is taken as

$$IVI_{[t_i,t_{i+1}]} = w * \left(atan\left( max\left(60 - MAP_{[t_i,t_{i+1}]}, 0\right) * \frac{\pi}{2w} \right)\right)^z * \frac{1}{DR_{[t_i,t_{i+1}]}} * (t_{i+1} - t_i).$$

Due to the limited occurrences of MAP values below 60, rather than being chosen based on a given data quantile, $w$ was taken equal to 14 as in the ‘excess’ vasopressor index case. The parameter $z$ was then selected to minimize, as above, the area under the curve of the squared difference between the function

$$x \to \ w * \left(atan\left(x * \frac{\pi}{2w}\right)\right)^z$$

and

$$x \to x$$

on $[0, w]$, resulting in $z = 1.6$.

Defining $IVI_{[t_i,t_{i+1}]}$with those values $w$ and $z$ guarantees that the increase in the 'insufficient' therapy variable is nearly linear as the MAP values decrease from 60 to 60-w=46. Additionally, it ensures that the 'insufficient' therapy variable remains at 0 for MAP values above 60. Figure E2 illustrates the effect of the non-linear transformation used in the 'insufficient' therapy index.

**Figure E2. Graphical representation of the effect of the non-linear transformation used in the ‘Insufficient’ therapy index.**

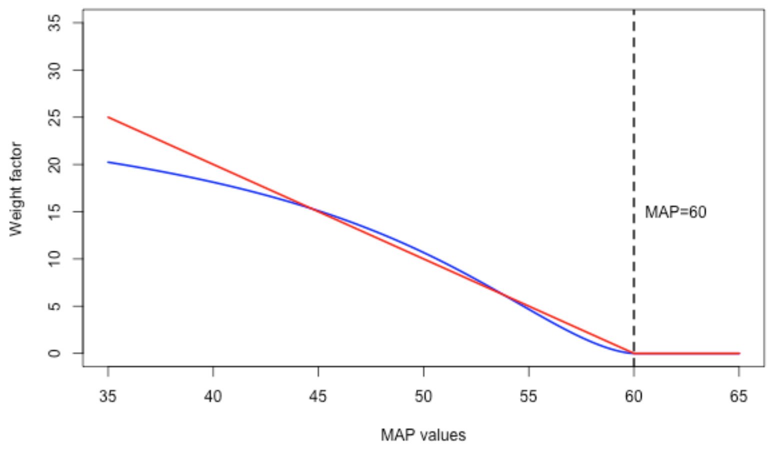

The blue curve represents the non-linear transformed values of the term $max(60 - MAP, 0)$ following the formula in equation (3). The red line represents the curve that is linear in the term $max(60 - MAP, 0)$ .

A patient's IVI is then defined as

$$IVI = \sum_{i=1}^{T-1} IVI_{[t_i, t_{i+1}]}$$

**Varying coefficient model**

The model equation can be expressed as

$$Logit\{P(Death)\} = \beta_0 + \beta_1(Apache)\ INDEX + \beta_2 Apache$$

where $\beta_1(Apache)$ is a logistic regression coefficient associated with the vasopressor exposure index variable considered, which is allowed to vary with respect to the Apache score variable. Here, it is suspected that the effect of excessive/insufficient dosage on mortality was weaker for patients with low severity scores at baseline (i.e., the "healthiest" among those admissible for the study) as well as for patients with high severity scores at baseline (i.e., the "sickest" among those admissible for the study), a quadratic formula for $\beta_1(Apache)$, i.e.,

$$\beta_1(Apache) = \beta_{10} + \beta_{11}\text{Apache} + \beta_{12}\text{Apache\^{}2}$$

The *varying coefficient model* names comes from the fact that, since the coefficient associated to the excess therapy variable can be interpreted as varying with respect to the Apache score variable.

When the equation for $\beta_1(Apache)$is plugged into the one for the logistic regression model above, one retrieves a standard logistic regression equation with two interaction terms:

$$Logit\{P(Death)\} = \beta_0 + \beta_{10}\,ET + \beta_2 Apache + \beta_{11} ET * \text{Apache} + \beta_{12} ET * \text{Apache\^{}2}$$

Because interaction terms have been incorporated into the model equation, the odds ratio of the excess therapy variable varies according to severity levels.

**Table E.1. Association between above and below MAP-vasopressor indices and hospital mortality (including all fitted coefficients)**

| | Derivation cohort | Validation cohort |
|---|---|---|
| Independent variables | OR (95% CI) | OR (95% CI) |
| Above MAP-vasopressor index | 1.6 (1.2 – 2.3) | 1.7 (0.9 – 4.0) |
| Adjustment variables | | |
| Age | 1.0 (1.0 – 1.1) | 1.0 (1.0 – 1.1) |
| APACHE II | 1.9 (1.4 – 2.5) | 2.6 (1.4 – 5.4) |
| Hypertension | 0.7 (0.4 – 1.2) | 2.2 (0.6 – 8.9) |
| Chronic dialysis | 0.4 (0.0 – 4.8) | 1.9 (0.2 – 26.9) |

| | | |
|---|---|---|
| Atherosclerosis | 0.4 (0.2 – 1.0) | 2.6 (0.7 – 10.5) |
| heart disease/heart failure | 0.4 (0.2 – 1.0) | 0.3 (0.0 – 2.5) |
| Sex (Female?) | 0.6 (0.3 – 1.0) | 3.8 (1.1 – 14.0) |
| Below MAP-vasopressor index | 1.1 (0.8 – 1.5) | 1.4 (0.7–2.6) |
| Adjustment variables | | |
| Age | 1.0 (1.0 – 1.1) | 1.0 (1.0–1.1) |
| APACHE II | 2.0 (1.5 – 2.7) | 3.4 (1.8–6.9) |
| Hypertension | 0.7 (0.4 – 1.2) | 1.5 (0.4–5.2) |
| Chronic dialysis | 0.2 (0.0 – 3.3) | 2.9 (0.3–47.5) |
| Atherosclerosis | 0.5 (0.2 – 1.3) | 2.4 (0.6–10.0) |
| heart disease/heart failure | 1.2 (0.6 – 2.3) | 0.2 (0.0–1.4) |
| Sex (Female?) | 0.6 (0.3 – 1.0) | 4.2 (1.3–15.4) |
